# Direct detection of microorganisms responsible for the sexually transmitted infection on the mobile real-time PCR device without treating in advance

**DOI:** 10.1101/2021.03.02.21252726

**Authors:** Masaaki Muraoka, Kazunori Sohma, Osamu Kawaguchi, Mikio Mizukoshi

**Affiliations:** Certified Non-Profit Organization Biomedical Science Association, 2-20-8, Kamiosaki, Shinagawa-ku, Tokyo, Japan

**Keywords:** sexually transmitted infection, *Chlamydia trachomatis*, *Neisseria gonorrhoeae*, *Treponema pallidum*, direct real-time PCR, rapid, quick, simple, easy, mobile

## Abstract

As WHO reported, four curable STIs-chlamydia, gonorrhoea, syphilis and trichomoniasis occur more than 1 million per each day globally almond 2016. For this reason, it is important to control these STIs, one of which is “to detect”. The general methods in order to detect STIs are nucleic acid amplification tests (NAATs). One of the reasons why NAATs are utilized in many tests is that it is possibly to be more sensitive than other test. However, there needs to treat extraction of nucleic acids in advance and amplify specific regions by NAATs, and hence it must take much labour and much time. In this work, for *Chlamydia trachomatis* (CT), *Neisseria gonorrhoeae* (NG) and *Treponema pallidum* (TP) which is each etiological agent of chlamydia, gonorrhoea and syphilis, we evaluate and propose “quicker and simpler” NAATs. Specifically, utilizing mobile real-time PCR device “PCR1100” and PCR reagent kit “KAPA3G Plant PCR Kit”, it was considered whether real-time direct PCR could be performed or not without treating DNA extraction in advance so-called “direct”.

As a result, firstly, we established that real-time direct PCR could be performed in all of CT, NG, and TP, and moreover, each Ct value correlated with the concentration of each organism similarly to detection of genome DNA (each correlation coefficient R^2^ > 0.95). Moreover, each assay demonstrated a limit of detection (LOD) of the follows; CT was 10^0.86 = 7.24 IFU/reaction, NG was 10^-0.19 = 0.65 CFU/reaction, and TP was 10^1.4 = 25.1 organisms/reaction. However, it appeared the sensitivity was a little low, especially for CT and TP.

Secondly, we found that even as without treating sample in advance, the time of detection was required more less 15 minutes at any of case, which was very quick compared with other current methods for real-time PCR. Additionally, compared with other commercial devices, it was easier to operate the PCR1100 device, for example, start, analysis of Ct value.

In conclusion, the present study has demonstrated that it is possible for real-time direct PCR to perform with combination of the PCR1100 device and the PCR reagent kit in 3 kinds of microorganisms-CT, NG and TP. Furthermore, we propose “quicker and simpler” methods for NAATs, which it would not take labour and time. Further studies are needed in order to contribute to control STIs.

## INTRODUCTION

According to the World Health Organization (WHO) global estimates for 2016, there were approximately 376 million for new infections of four curable STIs-chlamydia, gonorrhoea, syphilis and trichomoniasis; therefore, globally, more than 1 million of these STIs occur each day [1, 2]. These STIs cause acute urogenital conditions such as cervicitis, urethritis, vaginitis and genital ulceration. Chlamydia and gonorrhoea can cause serious short- and long-term complications, including pelvic inflammatory disease, ectopic pregnancy, infertility, chronic pelvic pain and arthritis. On the other hand, syphilis in adults, can cause neurological, cardiovascular and dermatological disease, and in pregnant, is the second leading cause of stillbirth globally, and moreover, results in prematurity, low birthweight, neonatal death, and infections in newborns [1, 3].

For chlamydia that etiological agent is *Chlamydia trachomatis* (CT), in 2016, new infection occurred with rate of 3.8 % females and 2.7 % males among 15–49 ages worldwide, especially with high rate in the African and Americas Regions [1, 2]. For gonorrhoea that etiological agent is *Neisseria gonorrhoeae* (NG), in 2016, new infection occurred with rate of 0.9 % females and 0.7 % males among 15–49 ages worldwide, and moreover, with the highest magnitude in the African Region. Co-infection with chlamydia is detected in 10–40 % of people with gonorrhoea [1, 4]. For syphilis that etiological agent is *Treponema pallidum* (TP), among reporting countries for 2019, an average of 3.2 % of antenatal care attendees were presumed to be positive [3]. Furthermore, an average of 10.8 % of sex workers tested were diagnosed with active syphilis [5]. STIs is spreading even now rather than the infection of the past as described, and moreover, the fact was also reported that almost half of incident STIs occurred in persons aged 15–24 years for 2019 [6]; therefore, it is very important to continue to control STIs in the future. In fact, the STIs control would be contributable to progress towards several Sustainable Development Goals (SDGs) related the health, such as “Goal 3: Good health and well-being” [7].

The detection of STIs is one of the important things as the STIs control, thus there have been a lot of methods in depending on specific substance. Throughout the last several decades, nucleic acid amplification tests (NAATs) have been developed. NAATs have the advantage of depending on nucleic acid materials only without requiring viable or intact organisms. For the diagnosis of chlamydia and gonorrhoea, especially, is generally more sensitive than culture method [8, 9, 10, 11]. For syphilis, NAATs have been utilized to examine specimens from any lesion exudate, tissue, or body fluid, and moreover, blood depending on syphilis stage [11, 12]. As seen from above, one of the reasons why NAATs are utilized in many tests is that it is possibly to be more sensitive than other test. However, there needs to treat extraction of nucleic acids in advance and amplify specific regions by NAATs, and hence it must take much labour and much time.

It was our object in this time to contribute the STIs control, one of which to improve NAATs method to classify STIs efficiently. Specifically, the goal of this study was to perform to construct that the system of real-time PCR could be utilized to detect each target organism for shorter time, moreover, without special treating DNA from samples in advance. As follows in this paper, we evaluate and propose “quicker and simpler” method, suitable for the actual occasion and useful to control STIs effectively.

## MATERIALS AND METHODS

### Preparation of each strain

#### *Chlamydia* strain

CT strain UW-36/Cx (ATCC® VR-886™) was purchased form American Type Culture Collection (ATCC, USA). It was received 1 mL as frozen in a vial, which titer was 7.3 x 10^6 IFU/mL. To perform to evaluate the study this time, it was thawed at 37 °C, of which, 50 μL was diluted with 450 μL of Dulbecco’s Phosphate Buffered Saline without Ca and Mg, liquid (D-PBS (-)) (Nacalai Tesque, Inc., Japan), so that 10-fold was prepared. In the same way, it was followed to prepare decimal serial dilutions.

#### *Neisseria* strain

NG was kindly provided by National Institute of Infectious Diseases (NIID, Japan). It was received as the preserved gelatin-disk, which titer was 6.45×10^6 CFU/disk. To perform to evaluate the study this time, it was dissolved per gelatin-disk with 1 ml of D-PBS (-) by mixing at 37 °C for 10 minutes. The dissolved suspension was the basis as undiluted, of which, 50 μL was diluted with 450 μL of D-PBS (-), so that 10-fold was prepared. In the same way, it was followed to prepare decimal serial dilutions.

#### *Treponema* strain

TP was kindly provided by Tokiwa Chemical Industries Co. Ltd. (Japan). It was received 0.2 mL per vial as frozen, which titer was 2.54 x 10^9 organisms/mL. To perform to evaluate the study this time, it was thawed at 37 °C, of which, 10 μL was diluted with 90 μL of D-PBS (-), so that 10-fold was prepared. In the same way, it was followed to prepare decimal serial dilutions.

#### DNA extraction

Each DNA was extracted from undiluted sample microbial by a commercial kit (QIAamp DNA Mini Kit, QIAGEN N.V., Netherlands). It was utilized 180 µL of thawed solution for CT, a sheet of gelatine-disk for NG, and 50 µL of thawed solution for TP. Following extraction, each extracted DNA was utilized to prepare decimal serial dilutions with TE buffer Solution (pH8.0, Nacalai Tesque, Inc., Japan).

#### Primer and Probe

To detect each microorganism by NAATs, we adopted the real-time PCR by hydrolysis probes commonly also referred to as TaqMan™. For CT and NG, the commercial kit (Chlamydia/Neisseria gonorrhoeae TaqMan Probe/Primer and Control Set, Norgen Biotek Corp., Canada) was utilized. This kit was contained the two kinds of probes, which one labelled with FAM fluorescent dye could detect CT, and the other labelled with Cy5 fluorescent dye could detect NG. Whereas, for TP, the other commercial kit (Treponema pallidum genesig® Detection Kit, Primerdesign™ Ltd., UK) was utilized, which contained the probe labelled with FAM fluorescent dye.

#### Real-time PCR

As previously reported [13], taking account of quickness and easiness, PCR in all of this trials were performed with mobile real-time PCR device PCR1100 (Nippon Sheet Glass Co. Ltd., Japan). KAPA3G Plant PCR Kit (Kapa Biosystems, Inc., United States) was utilized as the real-time PCR reagent of all trials in this, which it was showed the most adequate in all current commercial enzyme kits when preliminary comparative trials performed for the PCR1100 device (data not shown). As briefly describe the composition of PCR reagent, for CT and NG, 1 μL of a sample (extracted DNA samples or raw samples) was amplified in a 17 μL reaction solution containing 1x KAPA Plant PCR Buffer, 0.85 μL of 25mM MgCl_2_, 0.6 μL of 2.5 U/μL KAPA3G Plant DNA Polymerase (KAPA3G Plant PCR Kit), 2.0 μL of Chlamydia/Neisseria Primer & Probe Mix (Chlamydia/Neisseria gonorrhoeae TaqMan Probe/Primer and Control Set). Whereas, for TP, 1 μL of a sample (extracted DNA samples or raw samples) was amplified in a 17 μL reaction solution containing 1x KAPA Plant PCR Buffer, 0.85 μL of 25mM MgCl_2_, 0.6 μL of 2.5 U/μL KAPA3G Plant DNA Polymerase (KAPA3G Plant PCR Kit), 1.5 μL of TP specific primer / probe mix (Treponema pallidum genesig® Detection Kit). With preliminary tested, each composition was optimized by positive controls in each kit (data not shown).

The PCR condition applied in present work was programmed as follows: in CT and NG, the reaction was initially maintained at 95 °C for 15 seconds for enzyme activation, followed by 50 cycles of denaturation at 95 °C for 3.5 seconds and annealing simultaneous with extension at 60 °C for 15 seconds; in TP, the reaction was initially maintained at 95 °C for 15 seconds for enzyme activation, followed by 50 cycles of denaturation at 95 °C for 3.5 seconds and annealing simultaneous with extension at 61 °C for 9 seconds. With preliminary tested, each condition was optimized by positive controls in each kit (data not shown). Incidentally, the procedure of PCR1100 device was according to previously reported [14].

## RESULTS

### Evaluation with the extracted DNA

To begin with, when utilized this method including PCR1100 device, KAPA 3G Plant Kit, and commercial detection kit for CT, NG, and TP, it needed to evaluate whether correctly detecting or not. For this reason, DNA was extracted, purified, and concentrated from each of three kind of microbes followed by being utilized for the evaluation. Evidently from the data, it was possible to detect each of three kind of microbes, and moreover, each Ct value was quantitatively correlated with decimal serial dilutions (the correlation coefficient R^2^ > 0.99) (Fig. 1). These results indicated that this method could be applied to evaluate whether the unprocessed sample could be utilized or not. Moreover, the time of detection was required more less 15 minutes at any of case.

**Fig. 1.**
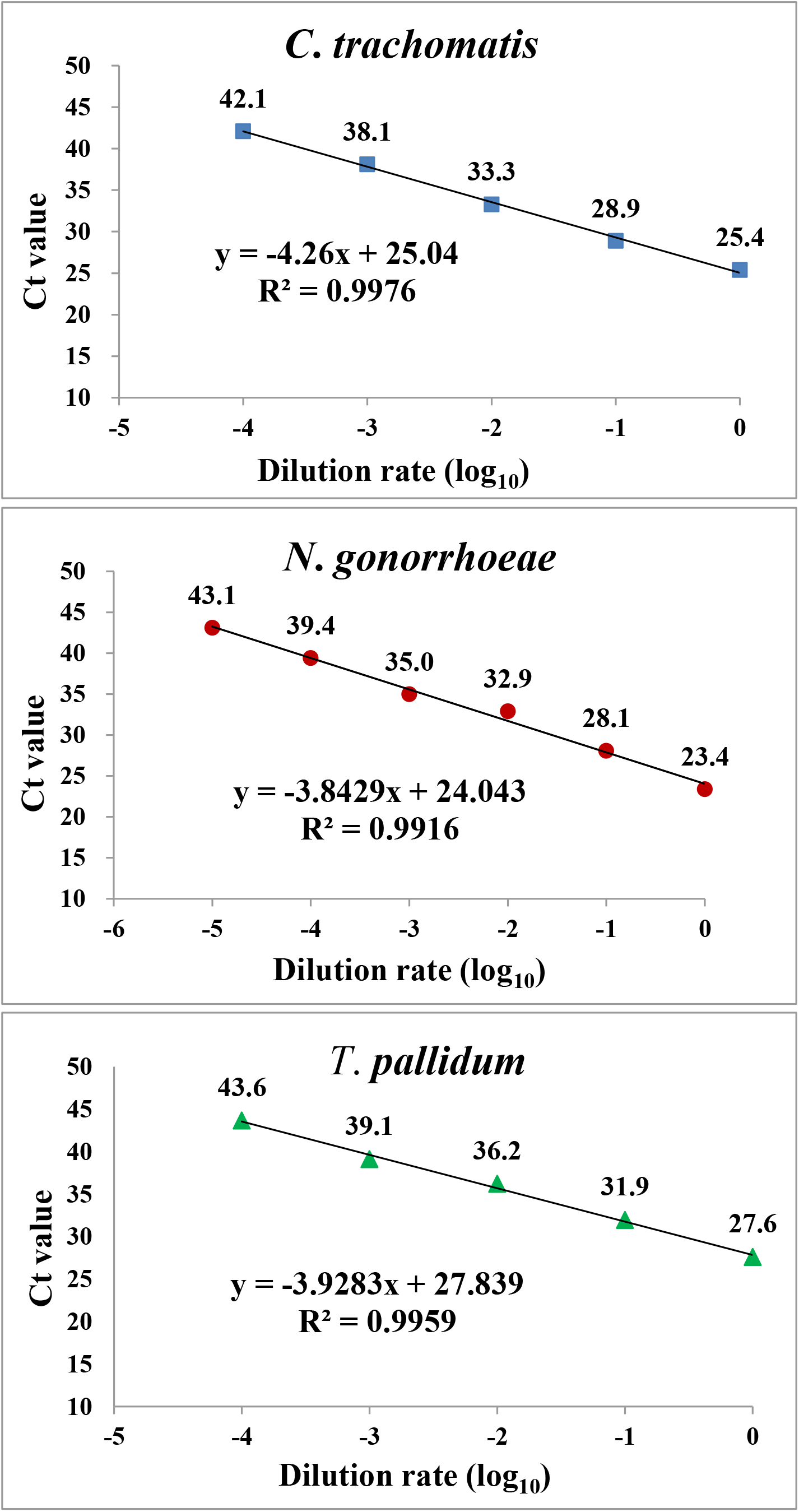
The correlation of each dilution and Ct value when the PCR1100 device utilized to detect extracted DNA. x: log_10_ Dilution rate; y: Ct value

### Detection in the raw sample

To close, even as without treating in advance such as extraction, purification and concentration of DNA, it evaluated whether correctly detecting each of three kind of microbes or not. Evidently from the data, it was possible to detect each of three kind of microbes, and moreover, each Ct value was quantitatively correlated with decimal serial dilutions (the correlation coefficient R^2^ > 0.95) (Fig. 2). Each assay demonstrated a limit of detection (LOD) of the follows; CT was 10^0.86 = 7.24 IFU/reaction, NG was 10^-0.19 = 0.65 CFU/reaction, and TP was 10^1.4 = 25.1 organisms/reaction. These results indicated that each of three kind of microbes could be directly detected by utilizing this method. Moreover, even as without treating in advance, the time of detection was required more less 15 minutes at any of case.

**Fig. 2.**
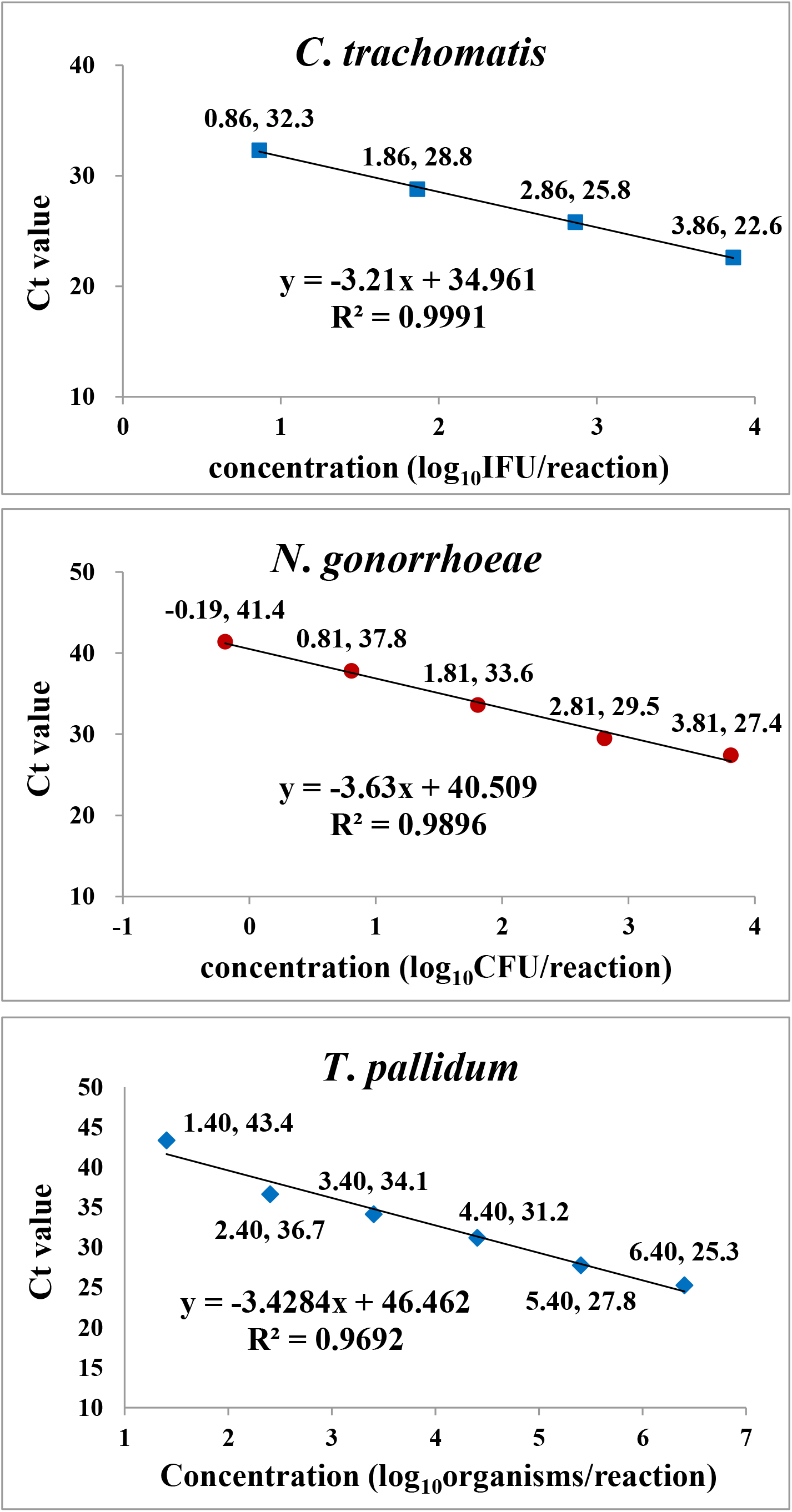
The correlation of each concentration and Ct value when the PCR1100 device utilized to detect raw sample. x: log_10_ Concentration; y: Ct value

## DISCUSSION

In this work, we had main objects to construct new methods for detection against each organism included in STIs by real-time PCR without extraction, concentration and purification of DNA from each sample, and moreover, to assess whether it was quick and simple to judge. Therefore, as previously reported [13], taking account of quickness and easiness, PCR in all of trials this time were performed with mobile real-time PCR device PCR1100 device. Simultaneously, KAPA3G Plant PCR Kit was utilized as the real-time PCR reagent of all trials in this since it was showed the most adequate for the PCR1100 device in all current commercial enzyme kits. Moreover, it was possible for this kit to perform direct PCR from crude plant samples such as leaf discs, seed samples, and other plant tissue types [15], hence it was considered whether other organisms could be utilized.

Firstly, it was necessary to determine the optimal method combined with the PCR1100 device about the commercial detection kit of each organism. Hence, each method was optimized by positive controls in each kit, followed by evaluating whether genome DNA extracted from each organism could be detected or not. As the result, combined with the PCR1100 device, it was proved that the kit selected was able to be utilized, furthermore, it was established that each Ct value correlated with the concentration of each genome DNA (each correlation coefficient R^2^ > 0.99) (Fig. 1). Accordingly, the optimal method was determined to utilize to assess subsequently. However, the concentration of each genome DNA was not measured, hence each LOD per concentration could not be determined this time. As soon as it was measured, each LOD must be determined, thereby, the sensitivity to each genome DNA could be revealed and compared with other real-time PCR devices.

In the next try, we assessed the method for real-time PCR without treating DNA in advance, so-called real-time direct PCR. As illustrated in Figure 2, it was found that real-time direct PCR could be performed in all of CT, NG, and TP, and moreover, each Ct value correlated with the concentration of each organism similarly to detection of genome DNA (each correlation coefficient R^2^ > 0.95). It was evident from these that when combined PCR1100 device with KAPA3G Plant PCR Kit, each raw sample could be detected with high correlation against each concentration as same as extracted genome DNA. Given the information of each LOD, however, it appeared the sensitivity was a little low, especially for CT and TP. As previously reported, CT count for swab samples was 450 IFU/100 μL from cervical specimens of women and 72 IFU/100 μL from urethral specimens of men [16], and the dark-field microscopy of the ulcers among methods utilized for direct detection of TP is relatively insensitive, which of sensitivity is approximately 10^5^ organisms/ml [17]. There are room for improvement, example for increase of sample volume. The improvement of sensitivity requires further investigation.

Meanwhile, even as without treating in advance, the time of detection was required more less 15 minutes at any of case, which was very quick compared with other current methods for real-time PCR. This work indicated that these methods reduced time to detection by up to from 50 to 80 per cent compared with existing methods [18, 19, 20, 21]. Additionally, compared with other commercial devices, it was easier to operate the PCR1100 device, for example, start, analysis of Ct value. Several literatures suggest that self-collection of samples for use with NAATs may increase uptake of screening and reduce the burden of clinical time [11, 22, 23]. Considering the evaluation this time, this method would help doing self-collection of samples followed by self-detection in the place, and hence could contribute to control STIs. However, when making it easy for anyone to do self-detection, it must be needed to create and provide the special reagent. Moreover, further studies are needed to yield any findings about directly detection, namely, it should be evaluated for each biopsy specimen such as cervicitis, urethritis, vaginitis and genital ulceration, and moreover, rectum and pharynx. As a result, each method in this work would contribute to control STIs.

Finally, in this work, it could not evaluate *Trichomonas vaginalis* (TV) that is etiological agent for trichomoniasis among four curable STIs. The 2016 global prevalence of trichomoniasis estimates were 5.3% in women and 0.6% in men, thus this cannot be ignored [1]. Although Trichomoniasis may cause an abnormal vaginal discharge in women and be responsible for few cases of non-gonococcal urethritis in men, it may be asymptomatic in at least 50% of women, and 70–80% of men [11, 24]. For detection of TG, NAATs offer the greatest flexibility in sample collection methods as well as the highest sensitivity of all available diagnostic methods [25, 26, 27]. Moreover, residual genital samples for diagnosis of CT and NG utilizing NAATs are appropriate for detection of TG similarly [28]. For these reasons, it is possible to detect four curable STIs with utilizing multiplex, simultaneously. It was our object in this time to contribute to control four curable STIs. For this reason, it was proposed to be possible for the new NAATs method to identify STIs efficiently, especially “quicker and simpler”. Simultaneously, further studies are needed in order to make progress sustainably.

## STUDY DESIGN

When each sample in this work was assessed in the real-time PCR (PCR1100, Nippon Sheet Glass Co. Ltd., Japan), a ten-fold dilution series of positive control DNA (Chlamydia/Neisseria gonorrhoeae TaqMan Probe/Primer and Control Set, Norgen Biotek Corp., Canada, and Treponema pallidum genesig® Detection Kit, Primerdesign™ Ltd., UK) was indicated with high relativity to Ct value on the mobile real-time PCR device PCR1100 (This correlation coefficient R^2^ > 0.99) the same as on the another device.

## Data Availability

The data that support the findings of this study are available from the corresponding author, upon reasonable request.

## ACKNOWLEDGEMENTS

We thank Shunsuke Sejima at Certified Non-Profit Organization Biomedical Science Association for technical comments, and Takashi Fukuzawa in an ex-employee at Nippon Sheet Glass Co. Ltd. for technical comments.

## AUTHOR CONTRIBUTIONS

M. Muraoka conceived, designed experiments, performed experiments, analysed the data and wrote the manuscript. K. Sohma performed experiments and analysed the date. O. Kawaguchi conceived and designed experiments. M. Mizukoshi conceived and provided specimens. All authors also participated in the editorial process and approved the manuscript.

## ETHICS STATEMENT

This study did not contain any studies involving human participants and animals performed by any of the authors.

## REFERRENCES

[1] J. Rowley, S. V. Hoorn, E. Korenromp, N. Low, M. Unemo, L. J. Abu-Raddad, R. M. Chico, A. Smolak, L. Newman, S. Gottlieb, S. S. Thwin, N. Broutet and M. M. Taylor, “Chlamydia, gonorrhoea, trichomoniasis and syphilis: global prevalence and incidence estimates, 2016,” Bulletin of the World Health Organization, vol. 97, no. 8, pp. 548-562P, 1 Aug 2019., doi: 10.2471/BLT.18.228486

[2] World Health Organization, “Four curable sexually transmitted infections still affect millions worldwide,” Updated 6 June 2019. Available from: https://www.who.int/news/item/06-06-2019-four-curable-sexually-transmitted-infections-still-affect-millions-worldwide.

[3] World Health Organization, “THE GLOBAL HEALTH OBSERVATORY, Syphilis in pregnancy,” Updated 2021. Available from: https://www.who.int/data/gho/data/themes/topics/indicator-groups/indicator-group-details/GHO/antenatal-care-(anc)-attendees-tested-for-syphilis.

[4] World Health Organization, “Target Product Profiles for improved antimicrobial stewardship for gonococcal infection,” Updated 1 Sep 2019. Available from: https://www.who.int/news/item/01-09-2019-target-product-profiles-for-improved-antimicrobial-stewardship-for-gonococcal-infection.

[5] World Health Organization, “THE GLOBAL HEALTH OBSERVATORY, Sex workers with active syphilis,” Updated 2021. Available from: https://www.who.int/data/gho/data/themes/topics/indicator-groups/indicator-group-details/GHO/sex-workers-with-active-syphilis.

[6] K. M. Kreisel, I. H. Spicknall, J. W. Gargano, F. M. Lewis, R. M. Lewis, L. E. Markowitz, H. Roberts, A. S. Johnson, R. Song, S. B. S. Cyr, E. J. Weston, E. A. Torrone and H. S. Weinstock, “Sexually Transmitted Infections Among US Women and Men: Prevalence and Incidence Estimates, 2018,” Sexually Transmitted Diseases, 23 Jan 2021., doi: 10.1097/OLQ.0000000000001355

[7] UNITED NATIONS, “Sustainable Development Goal 3: Good health and well-being,” Available from: https://www.un.org/sustainabledevelopment/health/

[8] L. H. Bachmann, R. E. Johnson, H. Cheng, L. Markowitz, J. R. Papp, F. J. Palella, Jr. and E. W. Hook 3rd, “Nucleic acid amplification tests for diagnosis of Neisseria gonorrhoeae and Chlamydia trachomatis rectal infections,” Journal of Clinical Microbiology, vol. 48, no. 5, p. 1827–1832, 24 Mar 2010., doi: 10.1128/JCM.02398-09

[9] M. J. Mimiaga, D. J. Helms, S. L. Reisner, C. Grasso, T. Bertrand, D. J. Mosure, H. Weinstock, C. McLean and K. H. Mayer, “Gonococcal, chlamydia, and syphilis infection positivity among MSM attending a large primary care clinic, Boston, 2003 to 2004,” Sexually Transmitted Diseases, vol. 36, no. 8, pp. 507–11, Aug 2009., doi: 10.1097/OLQ.0b013e3181a2ad98

[10] J. Schachter, J. Moncada, S. Liska, C. Shayevich and J. D. Klausner, “Nucleic acid amplification tests in the diagnosis of chlamydial and gonococcal infections of the oropharynx and rectum in men who have sex with men,” Sexually Transmitted Diseases, vol. 35, no. 7, pp. 637–42, Jul 2008., doi: 10.1097/OLQ.0b013e31817bdd7e

[11] World Health Organization, “Laboratory diagnosis of sexually transmitted infections, including human immunodeficiency virus,” 2013., ISBN: 978 92 4 150584 0

[12] A. Gayet-Ageron, S. Lautenschlager, B. Ninet, T. V. Perneger and C. Combescure, “Sensitivity, specificity and likelihood ratios of PCR in the diagnosis of syphilis: a systematic review and meta-analysis,” Sexually Transmitted Infections, vol. 89, no. 3, pp. 251–6, May 2013., doi: 10.1136/sextrans-2012-050622

[13] M. Muraoka, Y. Tanoi, T. Tada, M. Mizukoshi and O. Kawaguchi, “Quickly and simply detection for coronaviruses including SARS-CoV-2 on the mobile Real-Time PCR without treating RNA in advance,” medRxiv, 2020; preprint., doi: 10.1101/2020.08.06.20168294

[14] K. Shirato, N. Nao, S. matsuyama and T. Kageyama, “Ultra-Rapid Real-Time RT-PCR Method for Detecting Middle East Respiratory Syndrome Coronavirus Using a Mobile PCR Device,” Japanese Journal of Infectious Diseases, vol. 73, no. 3, pp. 181–186, 2020., doi: 10.7883/yoken.JJID.2019.400

[15] SelectScience 2021, “KAPA3G Plant PCR Kits by Kapa Biosystems, Inc.,” Updated 15 Oct 2020. Available from: https://www.selectscience.net/products/kapa3g-plant-pcr-kits/?prodID=203641#tab-2

[16] L. O. Eckert, R. J. Suchland, S. E. Hawes and W. E. Stamm, “Quantitative Chlamydia trachomatis cultures: correlation of chlamydial inclusion-forming units with serovar, age, sex, and race,” The Journal of Infectious Diseases, vol. 182, no. 2, p. 540–544, Aug 2000., doi: 10.1086/315738

[17] H. Liu, B. Rodes, C. Y. Chen and B. Steiner, “New Tests for Syphilis: Rational Design of a PCR Method for Detection of Treponema pallidum in Clinical Specimens Using Unique Regions of the DNA Polymerase I Gene,” Journal of Clinical Microbiology, vol. 39, no. 5, pp. 1941–6, May 2001., doi: 10.1128/JCM.39.5.1941-1946.2001

[18] M. J. Espy, J. R. Uhl, L. M. Sloan, S. P. Buckwalter, M. F. Jones, E. A. Vetter, J. D. Yao, N. L. Wengenack, J. E. Rosenblatt, F. R. Cockerill III and T. F. Smith, “Real-time PCR in clinical microbiology: applications for routine laboratory testing.,” CLINICAL MICROBIOLOGY REVIEWS, vol. 19, no. 1, p. 165–256., Jan. 2006., doi: 10.1128/CMR.19.1.165-256.2006

[19] K. Jaton, J. Bille and G. Greub, “A novel real-time PCR to detect Chlamydia trachomatis in first-void urine or genital swabs,” Journal of Medical Microbiology, vol. 55, no. 12, pp. 1667–1674, Dec 2006., doi: 10.1099/jmm.0.46675-0

[20] S. O. Hjelmevoll, M. E. Olsen, J. U. Ericson Sollid, H. Haaheim, M. Unemo and V. Skogen, “A Fast Real-Time Polymerase Chain Reaction Method for Sensitive and Specific Detection of the Neisseria gonorrhoeae porA Pseudogene,” the Journal of Molecular Diagnostics, vol. 8, no. 5, p. 574–581., Nov 2006., doi: 10.2353/jmoldx.2006.060024

[21] A. G. Koek, S. M. Bruisten, M. Dierdorp, A. P. van Dam and K. Templeton, “Specific and sensitive diagnosis of syphilis using a real-time PCR for Treponema pallidum,” Clinical Microbiology and Infection, vol. 12, no. 12, pp. 1233–6, Dec 2006., doi: 10.1111/j.1469-0691.2006.01566.x

[22] J. G. Rosenberger, B. Dodge, B. Van Der Pol, M. Reece, D. Herbenick and J. D. Fortenberry, “Reactions to Self-Sampling for Ano-Rectal Sexually Transmitted Infections Among Men Who Have Sex with Men: A Qualitative Study,” Archives of Sexual Behavior, vol. 40, no. 2, pp|p. 281-8, Apr 2011., doi: 10.1007/s10508-009-9569-4

[23] B. Dodge, B. Van Der Pol, J. G. Rosenberger, M. Reece, A. M. Roth, D. Herbenick and J. D. Fortenberry, “Field collection of rectal samples for sexually transmitted infection diagnostics among men who have sex with men,” International Journal of STD & AIDS, vol. 21, no. 4, pp. 260–4, Apr 2010., doi: 10.1258/ijsa.2009.009056

[24] A. C. Seña, W. C. Miller, M. M. Hobbs, J. R. Schwebke, P. A. Leone, H. Swygard, J. Atashili and M. S. Cohen, “Trichomonas vaginalis infection in male sexual partners implications for diagnosis, treatment, and prevention,” Clinical Infectious Diseases, vol. 44, no. 1, pp. 13-22, 1 Jan 2007., doi: 10.1086/511144

[25] J. A. Jordan, D. Lowery and M. Trucco, “TaqMan-based detection of Trichomonas vaginalis DNA from female genital specimens,” Journal of Clinical Microbiology, vol. 39, no. 11, pp. 3819–22, Nov 2001., doi: 10.1128/JCM.39.11.3819-3822.2001

[26] A. Pillay, F. Radebe, G. Fehler, Y. Htun and R. C. Ballard, “Comparison of a TaqMan-based real-time polymerase chain reaction with conventional tests for the detection of Trichomonas vaginalis,” Sexually Transmitted Infections, vol. 83, no. 2, pp. 126–9, Apr 2007., doi: 10.1136/sti.2006.022376

[27] J. Schirm, P. A. J. Bos, I. K. Roozeboom-Roelfsema, D. S. Luijt and L. V. Möller, “Trichomonas vaginalis detection using real-time TaqMan PCR,” Journal of Microbiological Methods, vol. 68, no. 2, pp. 243–7, Feb 2007., doi:10.1016/j.mimet.2006.08.002

[28] B. Van Der Pol, C. S. Kraft and J. A. Williams, “Use of an adaptation of a commercially available PCR assay aimed at diagnosis of chlamydia and gonorrhea to detect Trichomonas vaginalis in urogenital specimens,” Journal of Clinical Microbiology, vol. 44, no. 2, pp. 366–73, Feb 2006., doi: 10.1128/JCM.44.2.366-373.2006

